# Review and Meta-Analysis of BEST1 Retinopathy: Global Prevalence and Mutational Landscape

**DOI:** 10.64898/2026.01.15.26343613

**Authors:** Matthijs Leenders, Mathijs Gaastra, Sarah Tuller, George Magrath, Ash Jayagopal, Karen E. Malone

**Affiliations:** GeneScape, Leiden, The Netherlands; Opus Genetics, Durham, North Carolina, United States

## Abstract

**PURPOSE:** BEST1-associated inherited retinal disease constitutes one of the largest inherited retinal disease patient populations across the world. Innovative therapies are currently in development to address this significant unmet need. To better understand the scale of unmet need, and the distribution of phenotypes and genotypes, we conducted a meta-analysis of BEST1 patients reported in the literature to provide up-to-date patient number estimates.

**METHODS:** We utilized the GeneScape® *IRD Patient Atlas*, an expertly curated database of ∼73K retinal disease patients undergoing extensive genetic testing, in combination with a dedicated literature search to estimate the proportion of IRD patients attributed to *BEST1* using fixed effects weighting. This was also translated to patient number estimates for each country. Further extrapolation of patient subtypes was estimated based on cohorts of *BEST1* patients reporting phenotypes and genotypes. In addition, a summary of contributing variants is reported by region.

**RESULTS:** Across regions BEST1 retinopathy patients are estimated to occur from 1 in 38K (in Germany) to 1 in 363K in Japan. In the western countries, bi-allelic patients are expected to contribute nearly 20% of the total BEST1 population, whereas one third of patients in East Asia are anticipated to be bi-allelic.

**CONCLUSIONS:** While patients are reported across the world, their prevalence and composition vary across geographies. In the literature bi-allelic patients are less often reported explicitly as ARB patients in the United States as compared to Europe. Variant diversity is reflected in regional reports and drives phenotypic distribution.

## Introduction

BEST1-driven retinal diseases (bestrophinopathies) are caused by pathogenic variants in the *BEST1* gene and forms part of a heterogeneous spectrum of phenotypes within inherited retinal dystrophies (IRDs), a group of genetic disorders characterized by progressive retinal degeneration that ultimately leads to vision loss and blindness. The most commonly associated disease with *BEST1* is Best vitelliform macular dystrophy (BVMD, also known as Best’s Disease), which is the second most frequent form of macular dystrophy (MD) and the most frequent autosomal dominant macular dystrophy.^1,2^ In addition, patients may manifest with more aggressive forms of degeneration due to autosomal recessive Best disease (ARB) or autosomal dominant vitreoretinochoroidopathy (ADVIRC). The full range of diagnoses associated with *BEST1* mutations may also include retinitis pigmentosa, generalized macular dystrophy, other eye diseases, as well as more complex syndromes.^3^

The BEST1 protein forms homo-pentameric channels that can be found in the eye, as well as the kidney, the central nervous system, and the testis. It is an important ion channel implicated in normal ocular development and functions as a calcium-activated chloride channel, a large anion channel, and potentially a gamma-aminobutyric acid (GABA) and glutamate transport channel, among other purported functions. Over 250 variants in the *BEST1* gene have been implicated in disease and affect the BEST1 protein in multiple ways.^4,5^ Generally, variants resulting in no or little expression of BEST1 protein, also referred to as null-variants, are associated with autosomal recessive disease. In addition, several primarily missense variants have been shown to exert dominant negative effects on BEST1 channels and are also inherited in an autosomal dominant manner.^6^

Opportunities for treating BEST1 with genetic therapies are dependent on the underlying variant pathomechanisms. Autosomal recessive null-variants are strong candidates for gene augmentation therapy, while gain-of-function variants likely will require more precise therapies, such as CRISPR/Cas9 gene editing. While gene augmentation approaches were thought initially to be only appropriate for ARB, recent research has suggested that the vast majority of BVMD autosomal dominant variants are loss-of-function variants with respect to CaCC activity, and that functional restoration of BEST1 may be possible with gene therapies as those mutations may functionally resemble recessive variants.^7^, 8

To date, there are no approved treatment options available for any of the bestrophinopathies, but several innovative approaches are in development. Therefore, investigating the molecular disease mechanism of each variant could help to further inform the choice of treatment strategies, which currently holds a large potential to improve the therapeutic landscape of inherited retinal diseases.^9^

Still, the phenotypic and genetic heterogeneity, as well as overlapping features, pose a diagnostic challenge for defining the subsets of BEST1-driven disease. To support a most up-to-date patient view on BEST1-driven inherited retinal disease in the primary regions of interest, we conducted literature-based meta-analysis to capture the full range of BEST1-driven retinal disease. In addition, we summarized the most common variants identified in different geographies.

## Methods

### Estimating BEST1 Patient Prevalence Using Systematic Literature Review

To capture the breadth of BEST1-associated ophthalmic diagnoses, we conducted a systematic literature review for meta-analysis in accordance with PRISMA 2020 reporting guidelines. A structured search strategy was developed a priori and applied to PubMed, using predefined search terms (Supplementary Fig. 1).

All retrieved records underwent title and abstract screening by a single reviewer, using prespecified inclusion and exclusion criteria. Records were excluded if they were reviews, in vitro studies, small case reports, nonEnglish publications, studies without accessible full texts, or reports containing duplicate patient cohorts. In the latter case of suspected patient duplication, the report with the most comprehensive data reporting was considered. Fulltext articles were subsequently assessed for eligibility. After full-text review, reports were only included if they clearly stated the catchment, the total number of genetically tested IRD patients, and the number of patients attributed to *BEST1*, as this is the minimal information needed to conduct the meta-analysis. See Supplementary Fig. 1 for a summarized PRISMA flow diagram, including search-terms, inclusion criteria and an overview of papers identified.

Results of the dedicated PubMed search were supplemented with records from the GeneScape® *IRD Patient Atlas* that also met the inclusion criteria but not returned in the original search strategy. The I*RD Patient Atlas* is an expertly curated, literature-based database that aggregates data on IRD patients worldwide from publicly available sources. Hence, the *IRD Patient Atlas* integrates patient characteristics from diverse cohort studies, capturing phenotypic, demographic, and mutational information in a structured and accessible manner. The *IRD Patient Atlas* was queried for IRD cohorts in the target regions to identify appropriate datasets for estimating the proportion of IRD patients attributed to *BEST1* gene variants. Studies from the literature search or the *IRD Patient Atlas* were included for the meta-analysis ensuring no duplications. Some records in the *IRD Patient Atlas* were biased towards a clinical sub-group of IRDs (e.g. cohorts describing only patients with retinitis pigmentosa). These cohorts were excluded from this analysis. Notably, IRD cohorts sequencing the *BEST1* gene but without identifying any *BEST1* patients were also included to ensure robust estimates of the proportion. The final records included in the review for meta-analysis are found in Supplementrary Table S1.

After determining the *BEST1* proportion for each population using fixed effects weighting, we estimated the absolute number of probands affected by *BEST1*-related disease by projecting these proportions onto the total estimated IRD population for each country, as reported in the literature. The upper and lower limits of the estimates were determined with the modified Wald method. See Supplementary Table S2 for total IRD population estimates and related sources.

Since both the records from the systematic literature review and the literature-based *IRD Patient Atlas* have data consisting only of probands/index patients, we corrected for the probability of additional family members with dominantly inherited variants. This was done using the average fertility rate over the last 30 years for each country, obtained from *World Bank Open Data*^10^ (Supplementary Table 1), and the proportion of dominant *BEST1* variants for each country, including lower and upper limits. Prevalence was converted to annual incidence of new cases based on an age of onset of 26.8 years, as was recently reported in a large study of 222 cases in the United Kingdom.^2^

### Genotype-Phenotype Relations for BEST1

To estimate the genotype-phenotype relations for *BEST1* patients, we queried the *IRD Patient Atlas* to extract all *BEST1* patients in East Asia, Europe and the North America, extracting their phenotype and genotype, down to the variant level. See Supplemental Table S3 for all references. Using this information, the ratio of heterozygous (suggesting dominant inheritance of a variant) patients were determined. We could also determine relationships between the zygosity of *BEST1* patients and their clinical phenotype for each region. This was our final layer of epidemiological modelling. Calculated ratios for the relationships between region, zygosity, and clinical phenotypes were applied to the total estimated prevalence of *BEST1* patients in the regions by extrapolation.

### Determining the Mutational Spectrum of BEST1

We also evaluated the mutation landscape of *BEST1* in different regions: North America, Europe, and East Asia. This was done by querying the *IRD Patient Atlas* for all *BEST1* patients in the corresponding regions. Unsolved patients or patients with insufficient information on the variant level were excluded from this part of the analysis. From the remaining pool of patients for each region, all observed variants were analyzed for type and frequency. Variants were considered dominant negative or autosomal recessive if previously described in two extensive *BEST1*-variant studies.^4,5^ If the variant was not described in one of these studies, we looked for evidence of recessive inheritance in our dataset. The genetic variant was considered recessive if any of the following applied: (1) it was a nonsense, frameshift, or canonical splice-site variant, (2) homozygous cases were present in our dataset, or (3) it was observed among patients explicitly diagnosed with ARB in our dataset. For the remaining variants, we looked to see if there were heterozygous patients, suggesting autosomal dominant inheritance. The remaining variants were not classified as dominant or recessive. Topographic contour plots of the common recessive variants (p.Arg141His and p.Arg255Trp) were prepared using Surfer® from Golden Software, LLC (www.goldensoftware.com) based on geographically defined cohorts (see Supplementary Table 5).

## Results

*Identification and Inclusion of Eligible RecordsSupplemental Figure 1 shows a flow diagram, with the number of records used in each selection phases. In the screening phase, we excluded 142 records. In total, 27 research articles were included in the analysis that met the inclusion criteria. Supplementing these records withdata from the GeneScape® IRD Patient Atlas, we included a total of 65 records. Supplemental table 1 shows the records selected for the the BEST1 prevalence calculations for each region,*

**Figure 1.**
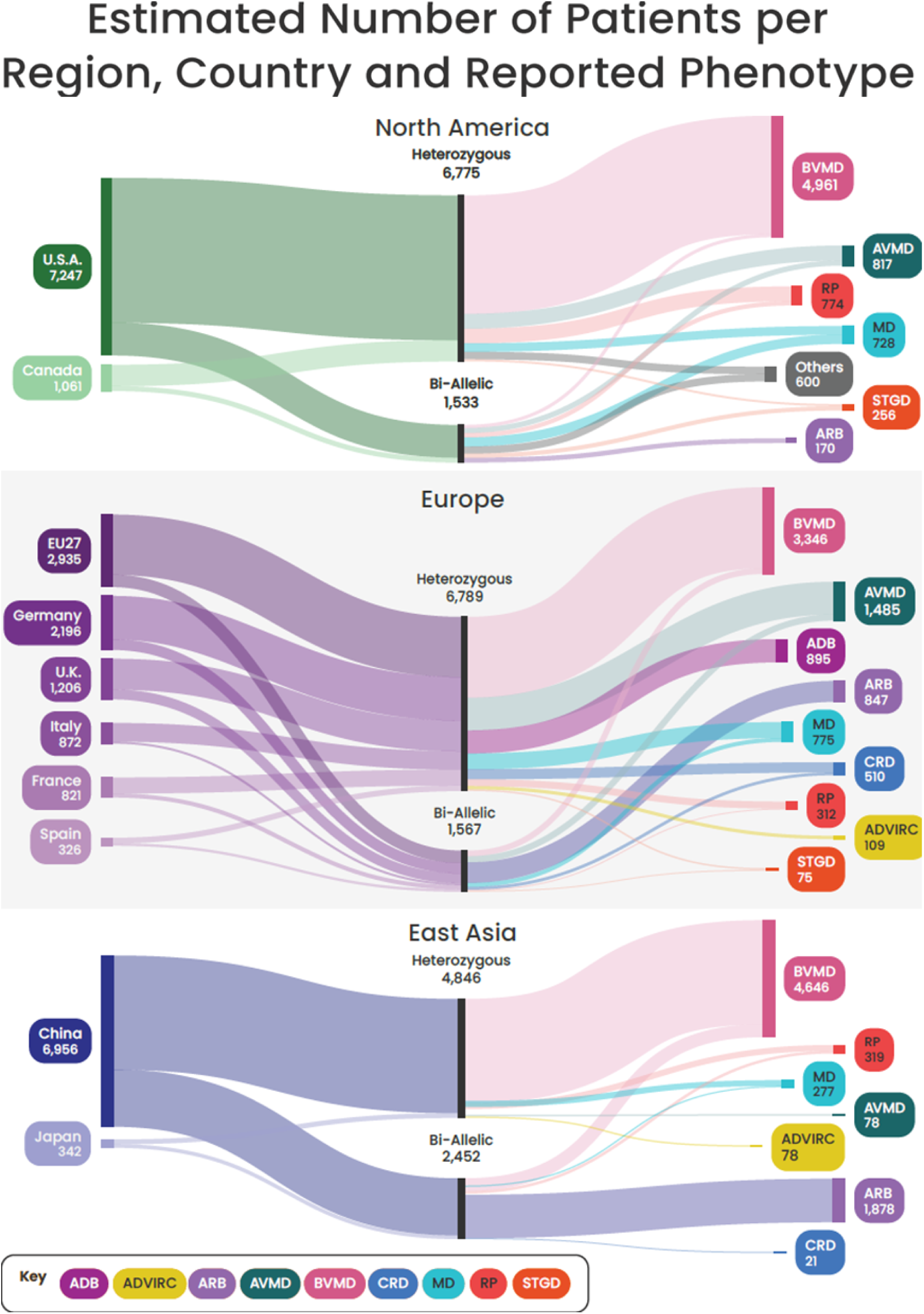
Estimated number of patients per region, country and reported phenotype. Sankey charts showing estimated patient numbers by genotype–phenotype relationships across regions and countries. Abbreviations: autosomal dominant bestrophinopathy (ADB); autosomal dominant vitreoretinochoroidopathy (ADVIRC); autosomal recessive bestrophinopathy (ARB); adult vitelliform macular dystrophy (AVMD); Best vitelliform macular dystrophy (BVMD); cone-rod dystrophy (CRD); macular dystrophy (MD); retinitis pigmentosa (RP); Stargardt disease (STGD). Note: EU27 refers to the remaining EU27 countries, not including France, Germany, Italy and Spain which are depicted independently.

### Estimated Prevalence of BEST1 Retinopathy Patients per Country

Using combined data from the systematic literature review and the IRD Patient Atlas, we estimated the number of retinal dystrophy patients attributed to *BEST1* variants across major regions (see Table 1). For the United States, we applied an IRD prevalence ratio of ∼1 in 1400.^11^ Based on our systematic review, IRD cohorts reported in the United States, 1.79% (95% confidence interval [CI], 1.60–2.01) had genetically confirmed BEST1 retinopathy, yielding an estimated 7247 patients (95% CI: 6434–8093) among the US population, after correcting for the probability of additional family members affected in dominant cases. Although no studies were specifically available for Canada, we applied the same proportions as we defined for the US and estimated that the total number of *BEST1* patients in Canada is ∼1061 (95% CI: 949-1194).

**Table 1.**
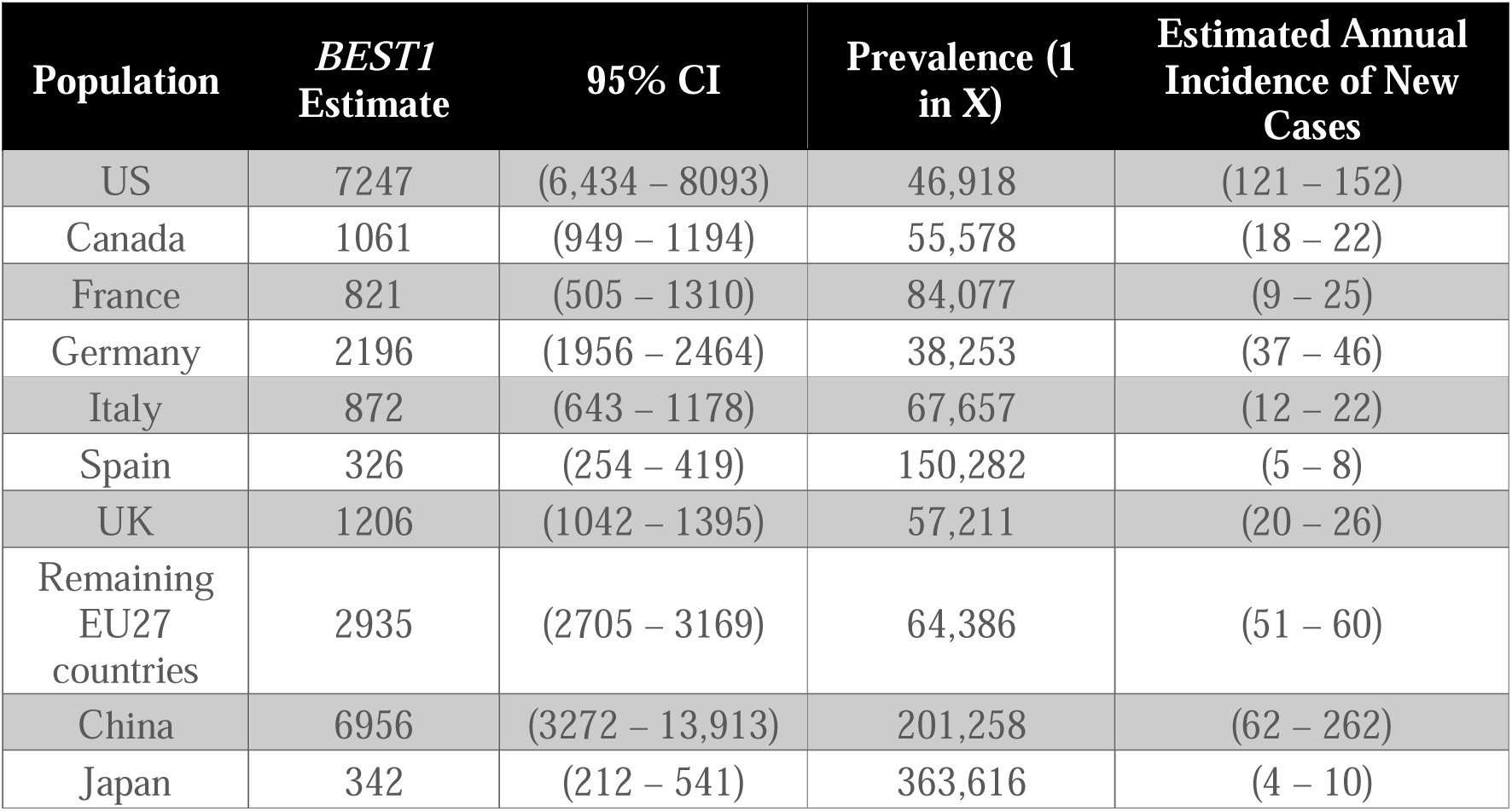
Estimated Prevalence and Incidence of *BEST1* Retinopathy Patients in Different Populations.

| Population | *BEST1*<br>Estimate | 95% CI | Prevalence (1<br>in X) | Estimated Annual<br>Incidence of New<br>Cases |
| --- | --- | --- | --- | --- |
| US | 7247 | (6,434 – 8093) | 46,918 | (121 – 152) |
| Canada | 1061 | (949 – 1194) | 55,578 | (18 – 22) |
| France | 821 | (505 – 1310) | 84,077 | (9 – 25) |
| Germany | 2196 | (1956 – 2464) | 38,253 | (37 – 46) |
| Italy | 872 | (643 – 1178) | 67,657 | (12 – 22) |
| Spain | 326 | (254 – 419) | 150,282 | (5 – 8) |
| UK | 1206 | (1042 – 1395) | 57,211 | (20 – 26) |
| Remaining<br>EU27<br>countries | 2935 | (2705 – 3169) | 64,386 | (51 – 60) |
| China | 6956 | (3272 – 13,913) | 201,258 | (62 – 262) |
| Japan | 342 | (212 – 541) | 363,616 | (4 – 10) |

In Europe, we analyzed approximately 28,000 IRD patients from 47 cohorts. Country-specific data for France, Germany, Italy, Spain, and the UK were used to make independent estimations for each of these countries (Table 1). Interestingly, the proportion of retinal disease patients attributed to *BEST1* in Spain (1.4%) and Italy (1.2%) was lower compared to the other three northwestern European countries (2.7% – 3.0%). Particularly in Spain, where the overall prevalence of IRDs is relatively low, the estimated number of individuals affected by BEST1-associated retinal disease was correspondingly limited. To calculate the remaining EU27 countries, all available European cohorts were utilized, including cohorts from countries such as Bulgaria, Denmark, Finland, Ireland, the Netherlands, Poland, Portugal, Slovenia, Sweden, and Switzerland. Across the entire EU27 countries, 2.27% (95% CI, 2.09–2.44) of IRD patients were *BEST1*-positive.

For East Asia, available data were limited. Based on data from 1559 IRD patients reported in Japan who underwent genetic testing for *BEST1*, we estimated that ∼342 *BEST1* cases can be found in Japan (95% CI: 212–541). In China, using a global IRD prevalence of 1 in 2500 (IRD population ∼560,000) and a cohort of 979 IRD patients undergoing *BEST1* genetic testing, we estimated ∼6956 cases (95% CI, 3272–13,913) may be found in China.

### BEST1-Driven Phenotypes in Different Regions

Using the *IRD Patient Atlas*, we can also estimate distribution of different *BEST1*-associated phenotypes. In addition to the large number of BEST1 patients identified in IRD cohorts, a substantial proportion are also reported in selected macular dystrophy^4,12^ and retinitis pigmentosa cohorts (see additional references in supplemental table 3).^13,14^ Figure 1 (top panel) illustrates the clinical phenotypes and their estimated prevalence in the US and Canada. Among 466 US *BEST1* probands with genotype-diagnosis level data in our virtual cohort, the majority are diagnosed with BVMD (Best disease), followed by AVMD, RP, and MD, with no ADVIRC cases identified. The middle panel of Figure 1 shows the corresponding distribution of European patients, derived from a virtual cohort of 1381 individuals with genotype-diagnosis data. Like the US, BVMD represents the largest subpopulation. Most bi-allelic genotypes are clearly classified as autosomal recessive Best disease in Europe. ADVIRC cases are also captured in the European dataset, though they represent a very small subgroup. For East Asia, the bottom panel of Figure 1 depicts the breakdown for Japan and China, derived from a virtual cohort of 161 patients.

### Summary of Observed BEST1 Variants Across Geographies

Complete genotypic information was available for subsets of the patients in the IRD Patient Atlas, enabling variant-level analysis. Figure 2 shows the genetic landscape of *BEST1* patients in the three different geographical regions. Variants associated with autosomal recessive Best disease (p.Arg141His, p.Ala195Val, p.Arg255Trp, p.Arg13His, and c.867+97G>A) are shown in lighter colors, and variants associated with dominant inheritance are shown in darker shade. For the U.S., 241 patients had detailed variant descriptions (Figure 2A), with the top four variants - all primarily associated with dominant inheritance - accounting for 25% of cases. In Europe, complete variant data were available for 1403 BEST1 patients. The most common variant in Europe was p.Ala243Val, associated with autosomal dominant BVMD, while p.Arg141His was the most frequent recessive variant (Figure 2B). Our East Asian cohort consisted of 434 patients from China, Japan, South Korea, and Taiwan. Here, the genetic landscape of BEST1 patients is largely driven by p.Arg255Trp, a mutation linked to autosomal recessive Best disease and originating in East Asian ancestry.^15^ Fig. 2B). Additionally, p.Ala195Val was identified as a relatively common variant worldwide.

**Figure 2.**
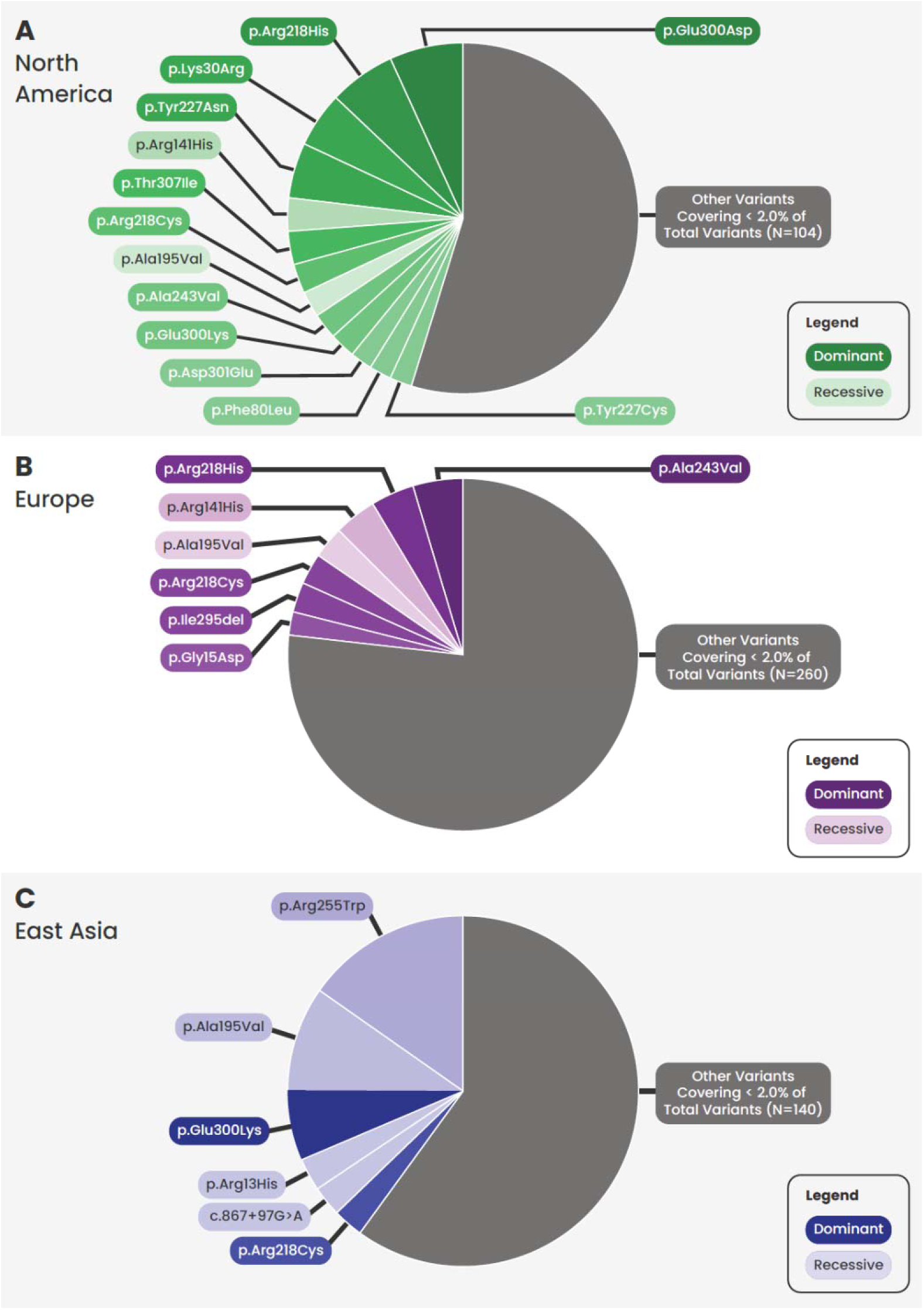
Mutational landscape reported per region. Pie chart depicting the contribution of *BEST1* variants. North America (A), Europe (B), and East Asia (C). Variants associated with autosomal recessive inheritance are shown in lighter colors, while darker colors indicate autosomal dominant inheritance.

Figure 3 presents the geographic distribution of the most common recessive variants observed in Europe (p.Arg141His) and Asia (p.Arg255Trp). Allele frequency data underlying these visualizations are provided in Supplementary Table S4. In Europe, p.Arg141His shows elevated frequencies in Scandinavian countries and the United Kingdom, suggesting a possible Nordic origin (Figure 3A). In contrast, Figure 3B highlights p.Arg255Trp in East Asia, where this variant exhibits its highest allele frequency among Korean and Northeastern Chinese populations, with notable occurrences among the Vietnamese population. Carriers for both variants are present in the U.S. (Figure 3C, Supplementary Table 5). p.Arg255Trp is predominantly associated with East Asian American ancestry, whereas most carriers of p.Arg141His are of European American descent.^16^

**Figure 3:**
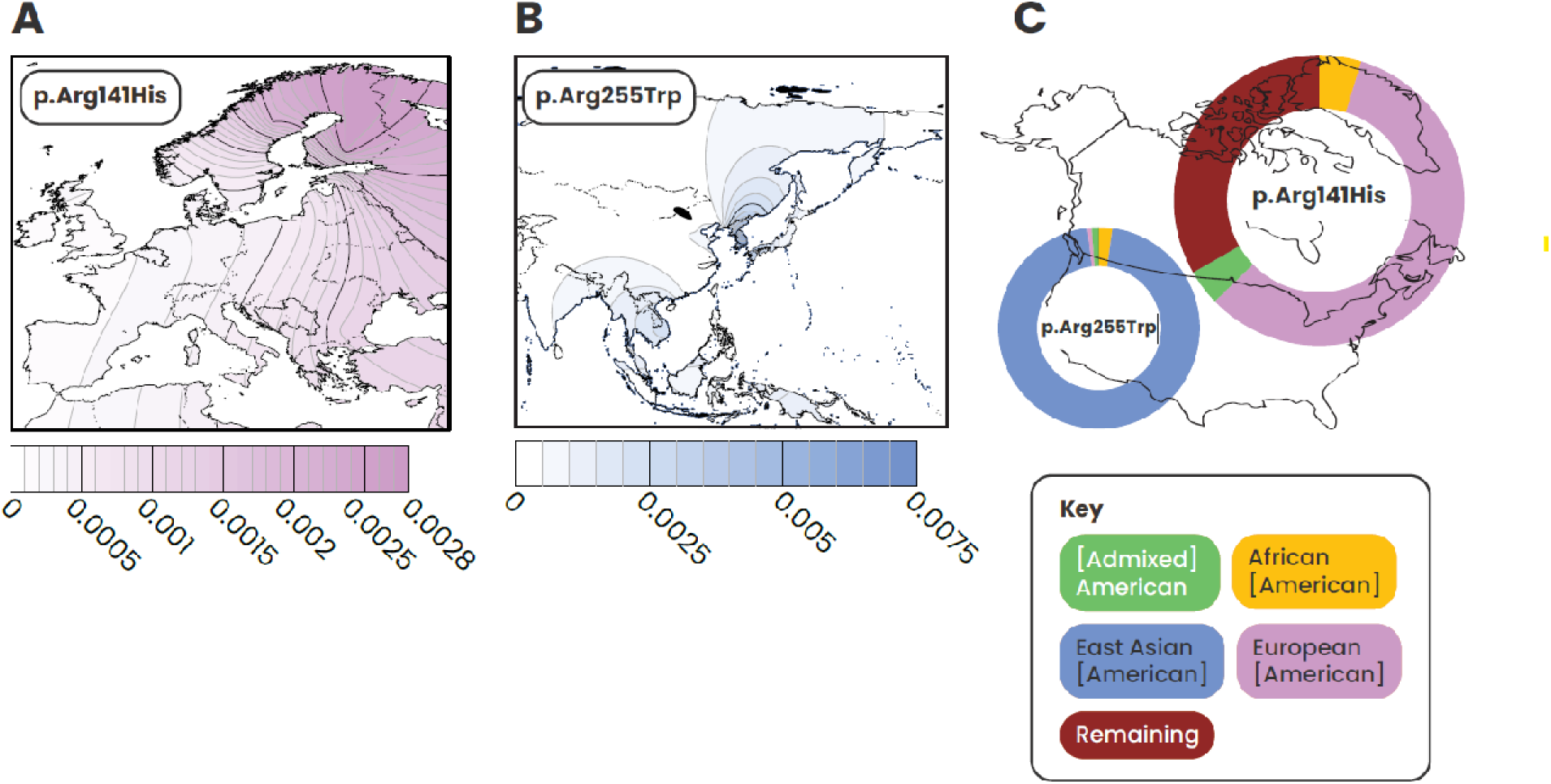
Inferred allelic frequencies across geographies for common autosomal recessive variants. Geographic distribution of common recessive *BEST1* variants: (A) p.Arg141His in Europe; (B) p.Arg255Trp in East Asia; (C) U.S. ancestry patterns from *All of Us*^16^ data, showing genetic ancestry contributions of p.Arg255Trp and p.Arg141His.

## Conclusion

In this study, we provide literature-based estimates of BEST1-driven retinopathy using the GeneScape *IRD Patient Atlas* combined with a dedicated literature search. Estimates for the number of patients for specific rare diseases have proved challenging^17^ but the increasing number of patients reported in the literature makes genetic epidemiology approaches for many rare diseases much more informative. Focusing on populations of the seven major markets, we determined that the current total disease prevalence is estimated between 11,072 and 15,434 patients, arising from mixed modes of inheritance. Further, we were able to derive the subtypes of BEST1 retinal disease and give population estimates for each target region. In North America and East Asia, a high number of bi-allelic patients do not appear to receive an ARB diagnosis and are more frequently captured under an umbrella diagnosis of Best disease. This discrepancy was less observed in European countries.

The mutational landscape of the *BEST1* gene differs in the regions assessed and drives the geographical differences in disease incidence and prevalence. We anticipate the most patients are in the U.S., however, European countries such as: Germany, the U.K., Italy, and France are expected to harbor some of the most concentrated populations of BEST1-driven retinal disease, including ARB subsets. Additionally, China is expected to harbor sizable BEST1 retinal disease populations, and notably the ARB subset is expected to compose nearly a third of the total Best disease population.

This analysis was specifically focused on BEST1-driven retinal disease populations in North America, Europe and East Asia. But it should also be noted that BEST1 patients are also reported in additional countries, including Australia, North Africa, the Middle East, and South American countries. The major limitations to this analysis are the bias in access and application of genetic testing, as well as uneven reporting in scientific literature across regions, where the United States and European countries are better represented. We also expect that these estimates may reflect an underestimation due to underdiagnosis, and lack of diagnosis generally observed in rare diseases.

These analyses represent the most up-to-date and comprehensive view of patient populations with BEST1-driven inherited retinal disease, including robust patient number estimates for regional prevalence and a clearer view of the most common variants driving disease across geographies. As innovative therapies advance through clinical stage development, identifying patients who could benefit from these therapies becomes increasingly important.

## Data Availability

All data produced in the present work are contained in the manuscript.

## Acknowledgements

We would like to thank Tarik Luisman for creating the Surfer plots in Figure 3.

## Commercial Relationships

M. Leenders, GeneScape (E), Opus Genetics (C); M. Gaastra, GeneScape (E) Opus Genetics (C); S. Tuller, Opus Genetics (E,I); A. Jayagopal, Opus Genetics (E,I); K.E. Malone, GeneScape (E), Opus Genetics (C)

## References

1. Rahman N, Georgiou M, Khan KN, Michaelides M. Macular dystrophies: clinical and imaging features, molecular genetics and therapeutic options. Br J Ophthalmol. 2020;104(4):451–460. doi:10.1136/bjophthalmol-2019-315086

2. Laich Y, Georgiou M, Fujinami K, et al. Best Vitelliform Macular Dystrophy Natural History Study Report 1: Clinical Features and Genetic Findings. Ophthalmology. 2024;131(7):845–854. doi:10.1016/j.ophtha.2024.01.027

3. Beryozkin A, Sher I, Ehrenberg M, et al. Best Disease: Global Mutations Review, Genotype-Phenotype Correlation, and Prevalence Analysis in the Israeli Population. Invest Ophthalmol Vis Sci. 2024;65(2):39. doi:10.1167/iovs.65.2.39

4. Kinnick TR, Mullins RF, Dev S, et al. Autosomal recessive vitelliform macular dystrophy in a large cohort of vitelliform macular dystrophy patients. Retina. 2011;31(3):581–595. doi:10.1097/IAE.0b013e318203ee60

5. Nachtigal AL, Milenkovic A, Brandl C, et al. Mutation-Dependent Pathomechanisms Determine the Phenotype in the Bestrophinopathies. Int J Mol Sci. 2020;21(5):1597. doi:10.3390/ijms21051597

6. Holtes LK, de Bruijn SE, Cremers FPM, Roosing S. Dual inheritance patterns: A spectrum of non-syndromic inherited retinal disease phenotypes with varying molecular mechanisms. Prog Retin Eye Res. 2025;104:101308. doi:10.1016/j.preteyeres.2024.101308

7. Sinha D, Steyer B, Shahi PK, et al. Human iPSC Modeling Reveals Mutation-Specific Responses to Gene Therapy in a Genotypically Diverse Dominant Maculopathy. Am J Hum Genet. 2020;107(2):278–292. doi:10.1016/j.ajhg.2020.06.011

8. Ji C, Li Y, Kittredge A, et al. Investigation and Restoration of BEST1 Activity in Patient-derived RPEs with Dominant Mutations. Sci Rep. 2019;9(1):19026. doi:10.1038/s41598-019-54892-7

9. Haldrup SB, McClements ME, Cehajic-Kapetanovic J, Corydon TJ, MacLaren RE. Gene Therapy Strategies for the Treatment of Bestrophinopathies. Int J Mol Sci. 2025;26(19). doi:10.3390/ijms26199421

10. Fertility rate, total (births per woman) | Data. Website. Accessed December 10, 2025. https://data.worldbank.org/indicator/SP.DYN.TFRT.IN

11. Abbass NJ, Yazji I, Allan KC, Kaelber DC, Talcott KE, Singh RP. Trends and Disparities in the Incidence and Prevalence of Inherited Retinal Diseases in the United States. Am J Ophthalmol. 2025;279:165–173. doi:10.1016/j.ajo.2025.07.021

12. Del Pozo-Valero M, Riveiro-Alvarez R, Martin-Merida I, et al. Impact of Next Generation Sequencing in Unraveling the Genetics of 1036 Spanish Families With Inherited Macular Dystrophies. Invest Ophthalmol Vis Sci. 2022;63(2):11. doi:10.1167/iovs.63.2.11

13. Koyanagi Y, Akiyama M, Nishiguchi KM, et al. Regional differences in genes and variants causing retinitis pigmentosa in Japan. Jpn J Ophthalmol. 2021;65(3):338–343. doi:10.1007/s10384-021-00824-w

14. Boulanger-Scemama E, El Shamieh S, Démontant V, et al. Next-generation sequencing applied to a large French cone and cone-rod dystrophy cohort: mutation spectrum and new genotype-phenotype correlation. Orphanet J Rare Dis. 2015;10(1). doi:10.1186/S13023-015-0300-3

15. Gao FJ, Gao FJ, Gao FJ, et al. Mutation spectrum of the bestrophin-1 gene in a large Chinese cohort with bestrophinopathy. British Journal of Ophthalmology. 2020;104(6):846–851. doi:10.1136/bjophthalmol-2019-314679

16. The “All of Us” Research Program. New England Journal of Medicine. 2019;381(7):668–676. doi:10.1056/NEJMSR1809937;WGROUP:STRING:MMS

17. Ferreira CR. The burden of rare diseases. Am J Med Genet A. 2019;179(6):885–892. doi:10.1002/ajmg.a.61124

