## Supplemental Materials for "Review and Meta-Analysis of BEST1 Retinopathy: Global Prevalence and Mutational Landscape"

Supplementary information: PRISMA search terms and definitions

Pubmed search terms and filters applied:

(((((((((((inherited retinal) AND (disease OR dystrophy)) OR (Adult-onset foveomacular vitelliform dystrophy)) OR (Autosomal dominant vitreoretinochoroidopathy)) OR (Autosomal recessive bestrophinopathy)) OR (Best vitelliform macular dystrophy)) OR (Retinitis pigmentosa)) OR (Hypopyon)) OR (Vitelliform)) OR (Vitelliruptive)) OR (bulls eye maculopathy)) AND (BEST1 OR bestrophin)

Filters: Humans

Search date: results until 20/10/2025

Records returned: 328

Inclusion Criteria: clearly stated (1) the catchment, the (2) total number of genetically tested IRD patients, AND the (3) number of patients attributed to BEST1

Supplemental Figure 1. Overview of search results – PRISMA Flow chart

   
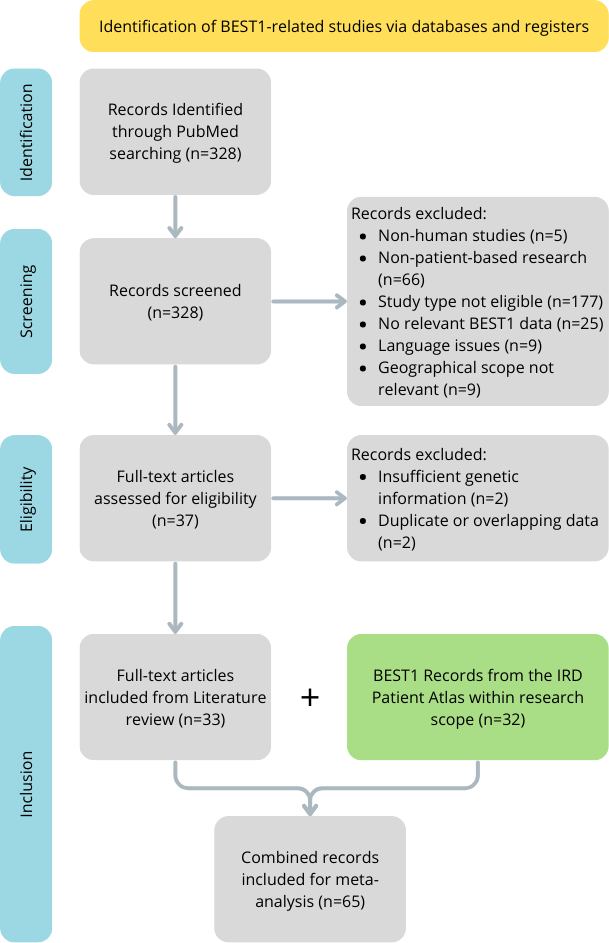

Supplemental Table S1. References (PMID) meeting the minimum inclusion criteria based on systematic PRISMA review for the BEST1 prevalence calculations for North America, East Asia and Europe

| North America | East Asia | Europe | | |
| --- | --- | --- | --- | --- |
| 25412400 | 25356976 | 22968130 | 31087526 | 34440443 |
| 28005406 | 26161267 | 23350551 | 31836858 | 34662339 |
| 28559085 | 33090715 | 23591405 | 31963381 | 34795310 |
| 29212538 | 36284460 | 23662838 | 32036094 | 35055178 |
| 29343940 |  | 26766544 | 32244552 | 35456422 |
| 31816670 |  | 27032803 | 32423767 | 35656873 |
| 32037395 |  | 27160483 | 32483926 | 36084042 |
| 32783387 |  | 27353947 | 32531858 | 36460718 |
| 32893963 |  | 27391102 | 32543920 | 36909829 |
| 34662339 |  | 27624628 | 33452396 | 37510321 |
| 34906470 |  | 28041643 | 33546218 | 37734845 |
| 35672425 |  | 28127548 | 33727790 | 38872169 |
| 36672815 |  | 28181551 | 33749171 | 38927562 |
| 39858579 |  | 28224992 | 33851411 | 38927702 |
| 40877827 |  | 30653986 | 34321860 | 39462066 |
|  |  | 30718709 | 34327195 |  |

Supplemental Table S2. Estimates of total IRD patients per country and related references. The fertility rate used is an average over the years 1994 – 2024.

| Country/Region | Total Estimated  IRD population | Fertility rate | Relevant References (PMID) or Registry |
| --- | --- | --- | --- |
| USA | 238063 | 1.91 | 40706695 |
| UK | 36365 | 1.74 | 24525390, UKBB |
| Spain | 18003 | 1.26 | 33727790,33452396,39277603 |
| France | 16870 | 1.89 | BNMDR (National Rare Disease Registry) |
| Germany | 45823 | 1.41 | 37606831 |
| Italy | 40180 | 1.31 | RNMR (National Rare Disease Registry) |
| Japan | 20854 | 1.36 | 39078460 |
| Canada | 35000 | 1.56 |  |
| China | 560000 | 1.56 |  |
| Remaining EU27 | 80900 | 1.53 |  |

Supplemental Table 3. References (PMID) used for BEST1 phenotype-genotype correlations for North America, East Asia, and Europe

| North America | Asia | Europe |  |
| --- | --- | --- | --- |
| 25097241 | 25324289 | 40891781 | 38927702 |
| 25412400 | 30498755 | 23350551 | 27353947 |
| 28005406 | 33090715 | 34240658 | 33452396 |
| 28559085 | 37747403 | 33529788 | 32543920 |
| 31736247 | 25489231 | 32531858 | 32036094 |
| 32783387 | 33691693 | 29847639 | 35119454 |
| 32893963 | 38499336 | 31963381 | 39189993 |
| 35672425 | 30593719 | 39596324 | 38872169 |
| 36672815 | 36378562 | 23591405 | 35456422 |
| 38347443 | 33629268 | 28224992 | 34795310 |
| 39858579 | 39048936 | 36460718 | 29555955 |
| 40877827 | 31144483 | 32423767 | 39462066 |
|  | 38195571 | 36909829 | 37734845 |
|  | 31816670 | 33749171 | 28041643 |
|  | 32100970 | 36819107 | 31570112 |
|  | 36284460 | 21412020 | 33727790 |
|  | 33608557 | 26103963 | 21738390 |
|  |  | 38700873 | 38540785 |
|  |  | 30718709 | 28127548 |
|  |  | 26766544 | 34327195 |
|  |  | 28181551 | 22334370 |
|  |  | 27624628 |  |

Supplemental Table 4. References (PMID) used for BEST1 variant-level analysis for North America, Europe and East Asia

| North America | East Asia | Europe |  |
| --- | --- | --- | --- |
| 21273940 | 25324289 | 21412020 | 33749171 |
| 25097241 | 25489231 | 21738390 | 34240658 |
| 25412400 | 30498755 | 22334370 | 34321860 |
| 28559085 | 30498755 | 23591405 | 34327195 |
| 31736247 | 30593719 | 26103963 | 34795310 |
| 32037395 | 31144483 | 26766544 | 35119454 |
| 34662339 | 31519547 | 27353947 | 35456422 |
| 35672425 | 31816670 | 27391102 | 35656873 |
| 36672815 | 32100970 | 28041643 | 36819107 |
| 38347443 | 33608557 | 28127548 | 36909829 |
|  | 33691693 | 28181551 | 37734845 |
|  | 36284460 | 28224992 | 38540785 |
|  | 36378562 | 29555955 | 38927562 |
|  | 37747403 | 30718709 | 38927702 |
|  | 38195571 | 31570112 | 39189993 |
|  | 38499336 | 32036094 | 39462066 |
|  | 39048936 | 32531858 | 40891781 |
|  |  | 33529788 |  |

Supplemental Table 5. Allele Frequency data for p.Arg255Trp and p.Arg141His

| **Location/Region** | **Source** | **AF_p.Arg255Trp** | **AF_p.Arg141His** |
| --- | --- | --- | --- |
| Beijing, China | Han100K | 0 | 0 |
| Bulgaria | gnomad_V2 | 0 | 0.001121 |
| Estonia | gnomad_V2 | 0 | 0.00105 |
| Finland | gnomad_V4 | 0 | 0.002608 |
| Henan, China | Han100K | 0 | 0 |
| NorthWestern Europe | gnomad_V2 | 0 | 0.000236 |
| Northern Sweden (Vasterbotten County) | ACPOP | 0 | 0.002 |
| Shanghai, China | Han100K | 0 | 0 |
| Southern Europe | gnomad_V2 | 0 | 0.000545 |
| Sweden | gnomad_V2 | 0 | 0.000559 |
| United Kingdom | UKBB | 0 | 0.00018 |
| Singapore | Han100K | 0.000183 | 0 |
| Liaoning, China | Han100K | 0.00036 | 0 |
| Guangdong, China | Han100K | 0.000378 | 0 |
| Jiangsu, China | Han100K | 0.000407 | 0 |
| Japan | TogoVar | 0.000543 | 0 |
| Hebei, China | Han100K | 0.0006 | 0 |
| South Korea | KOVA_2 | 0.000761 | 0 |
| Tianjin, China | Han100K | 0.000956 | 0 |
| Guangxi, China | Han100K | 0.001029 | 0 |
| Vietnam | Vietnamese Genetic Variation Database | 0.0025 | 0 |
| South Korea | GenomeAsia 100K | 0.0066 | 0 |

Supplemental Table 6. Allele Frequency (AF) of p.Arg255Trp and p.Arg141His among U.S. populations (All of Us)

|  | **AF_p.Arg255Trp** | **AF_p.Arg141His** |
| --- | --- | --- |
| African [American] | 0.000013 | 0.000025 |
| East Asian [American] | 0.000583 | 0 |
| European [American] | 0.000004 | 0.000307 |
| [Admixed] Americas | 0.000007 | 0.000021 |
| Middle Eastern [American] | 0 | 0 |
| American South Asian | 0 | 0 |
| Remaining | 0 | 0.000176 |
| Overall Allele Frequency | 0.000019 | 0.000184 |
